# Prediction of vaccine efficacy of the Delta variant

**DOI:** 10.1101/2021.08.26.21262699

**Authors:** Xinhua Chen, Andrew S. Azman, Wanying Lu, Ruijia Sun, Nan Zheng, Shijia Ge, Xiaowei Deng, Juan Yang, Daniel T. Leung, Hongjie Yu

## Abstract

The emergence of SARS-CoV-2 variants have raised concerns over the protective efficacy of the current generation of vaccines, and it remains unclear to what extent, if any, different variants impact the efficacy and effectiveness of various SARS-CoV-2 vaccines. We systematically searched for studies of SARS-CoV-2 vaccine efficacy and effectiveness, as well as neutralization data for variants, and used a previously published statistical model to predict vaccine efficacy against variants. Overall, we estimate the efficacy of mRNA-1273 and ChAdOx1 nCoV-19 against infection caused by the Delta variant to be 25-50% lower than that of prototype strains. The predicted efficacy against symptomatic illness of the mRNA vaccines BNT162b2 and mRNA-1273 are 95.1% (UI: 88.4-98.1%) and 80.8% (60.7-92.3%), respectively, which are higher than that of adenovirus-vector vaccines Ad26.COV2.S (44.8%, UI: 29.8-60.1%) and ChAdOx1 nCoV-19 (41.1%, 19.8-62.8%). Taken together, these results suggest that the development of more effective vaccine strategies against the Delta variant may be needed. Finally, the use of neutralizing antibody titers to predict efficacy against variants provides an additional tool for public health decision making, as new variants continue to emerge.

## Main

The coronavirus disease 2019 (COVID-19) pandemic, caused by severe acute respiratory syndrome coronavirus 2 (SARS-CoV-2) infection, was first reported in Wuhan, China, in late 2019, which had caused more than 4 million deaths globally and brought widespread social and economic disruption.^1^ It is widely accepted that the development of safe and effective COVID-19 vaccines would be the key to help bring the world to its pre-pandemic normalcy.^2^ As a result, there was a dramatic acceleration of vaccine development, with nearly 300 COVID-19 vaccine candidates in both clinical and pre-clinical development as of mid-2021.^3,4^ However, immune evasion caused by evolution and mutations of SARS-CoV-2 casts a shadow over the protective efficacy of existing licensed vaccines, which were developed based on prototype virus strains.^5,6^ Efficacy against the Delta variant, the current predominant circulating strain, is still unknown for the majority of licensed vaccines, and may be difficult to broadly ascertain, given the extensive resources required to identify and distinguish variants in vaccine trials. Although *Cromer et al* predicted variant-specific efficacy against symptomatic and severe infection, they didn’t estimate efficacy by different licensed vaccines and vaccine protection against SARS-CoV-2 infection.^7^ Here, we use statistical models to predict vaccine-specific efficacy against different SARS-CoV-2 variants and clinical endpoints, mainly for the Delta variant, as well as other variants of concern (VOCs) defined by the World Health Organization (WHO), across different clinical endpoints, using the previously established relationship between neutralization titer and protective efficacy, combined with *in vitro* cross neutralization assay results.

We conducted a systematic search from three peer-reviewed databases (PubMed, Web of Science and Embase) and an open science platform (Europe PMC) to included studies that are original analyses of COVID-19 vaccine efficacy with a randomized clinical trial design against wild type and variants, or are original analyses of COVID-19 vaccine effectiveness. We also conducted a systematic search to update a previously-reported meta-analysis of *in vitro* neutralization titers of individuals who have been vaccinated with prototype-strain-based vaccines against both SARS-CoV-2 prototype strains and variants.^8^ Combining these two datasets, we predicted vaccine protection against variants, following the statistical approach of *Khoury et al*, which used the relationship between neutralizing antibody levels and vaccine efficacy, as well as the fold-change of neutralizing antibodies titers against specific variants compared to prototype strain ^*9*^. We calculated uncertainty intervals of predicted efficacy by considering the 95% confidence interval limits from both fold-change of neutralizing titers and vaccine efficacy estimates against prototype strains. A detailed methodology is shown in Supplements.

According to the predefined search strategy, we systematically collected efficacy data against infection, symptomatic infection, and severe infection, with either prototype or mutant strains from a total of nine vaccines across four platforms **(Table S3)**. After matching efficacy data to neutralization data, the predicted efficacy of six vaccines can be estimated (BNT162b2, mRNA-1273, Gam-COVID-Vac, Ad26.COV2.S, ChAdOx1 nCoV-19, NVX-CoV2373), along with age-specific estimates for BNT162b2 and mRNA-1273. We found that the predicted efficacies against SARS-CoV-2 infection by the Delta variant are lower than that against reference lineage.

The predicted efficacy against infection of mRNA-1273 is 71.4% (UI: 62.3-79.1%) for the Delta variant, approximately 20% lower than predicted against the reference lineage. Similarly, the predicted efficacy for ChAdOx1 nCoV-19 against the Delta variant was 27.5% (UI: 12.4-46.0%), approximately 50% lower than predicted against the reference lineage **(Figure 1A)**. The predicted efficacies against Beta-variant-infection also showed a large reduction compared to prototype lineage, while predicted efficacies against the Alpha and Gamma were not reduced **(Figure S1)**.

**Figure 1.**
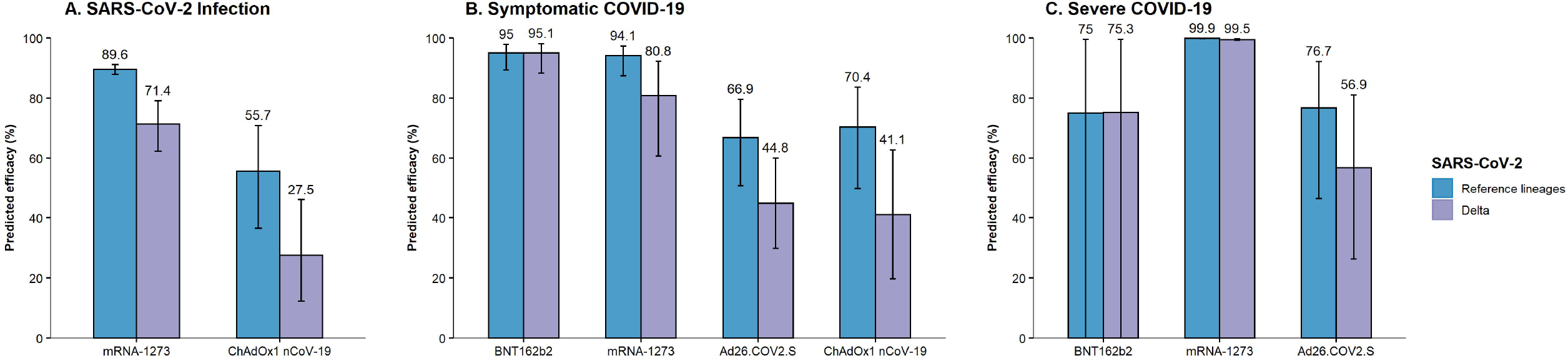
Predicted efficacy against the Delta variant across clinical endpoints. **A)** SARS-CoV-2 infection, **B)** Symptomatic COVID-19 and **C)** Severe COVID-19. The figure on the top of the bar represents the point estimate of the predicted efficacy. The error bar represents the uncertainty interval of predicted efficacy.

For the vaccine protection against symptomatic disease associated with the Delta variant, the predicted efficacies of two mRNA-based vaccines, BNT162b2 and mRNA-1273, are 95.1% (UI: 88.4-98.1%) and 80.8% (UI: 60.7-92.3%), respectively, compared to their prototype strain efficacy of approximately 95% **(Figure 1B)**. Age-specific efficacy for BNT162b2 was also largely retained compared to that of prototype strains **(Figure S2)**. The predicted efficacy of the two adenovirus-vector vaccines studied against the Delta variants are generally lower than that of mRNA vaccines and lower than that against prototype strain infection (range: 60%-70%), with 44.8% (UI: 29.8-60.1%) and 41.1% (UI: 19.8-62.8%) for Ad26.COV2.S and ChAdOx1 nCoV-19, respectively **(Figure 1B)**. For the other VOCs, the predicted efficacy against symptomatic disease was lower for Beta variant infection compared to other variants across all vaccines **(Figure S3)**.

We found that mRNA-1273 was predicted to provide nearly perfect protection against severe COVID-19 caused by the Delta variant **(Figure 1C)**, while the efficacies (against severe disease) of Ad26.COV2.S and BNT162b2 were predicted to be 56.9% (UI 26.3-81.1%), and 75.3% (UI 0.1-99.6%), respectively, though the latter had extremely broad uncertainty intervals due to the wide confidence intervals on vaccine efficacy against the prototypical strain **(Figure 1C)**.

When comparing to efficacy and effectiveness in published data, we find that most of the predicted efficacy correlated well with the vaccine efficacy from clinical trials and effectiveness in cohort/test-negative studies in the real world **(Figure 2)**. The predicted efficacy against symptomatic cases infected with the Delta variant for BNT162b2 (95.1; UI: 88.4-98.1) are generally consistent with the effectiveness data in United Kingdom (88.0%; 95%CI: 85.3-90.1%).^10^ A study in Canada reported that the effectiveness against symptomatic illness caused by the Delta variant for mRNA-1273 is 72% (95%CI: 57-82%),^11^ consistent with the predicted efficacy (80.8, UI: 60.7-92.3%) from this study **(Figure 2)**.

**Figure 2.**
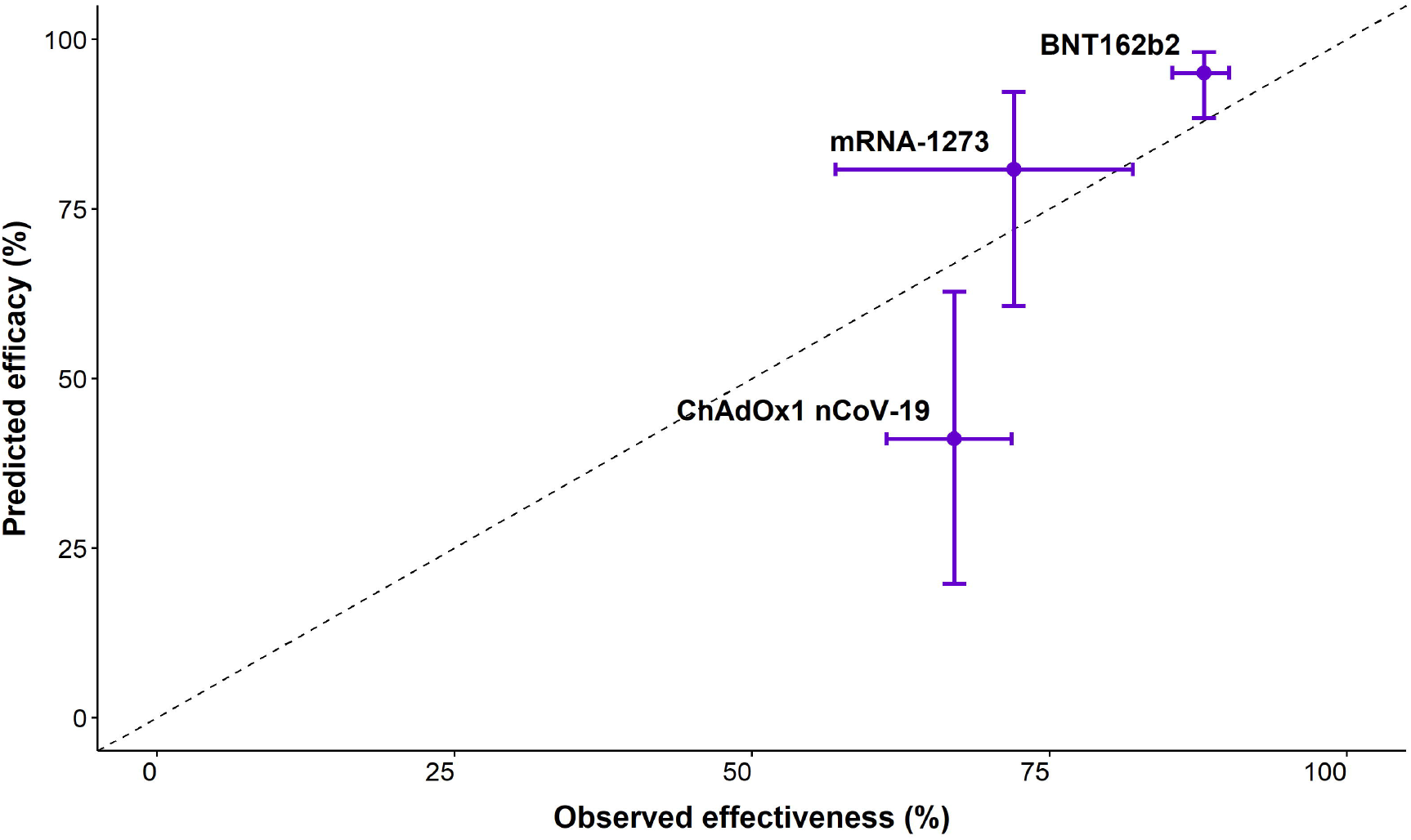
Comparison of predicted efficacy and observed effectiveness against symptomatic COVID-19. The vertical error bar represents the uncertainty interval of predicted efficacy. The horizontal error bar represents confidence interval of reported effectiveness.

Overall, we predicted COVID-19 vaccine efficacy against SARS-CoV-2 variants, especially for the current predominant circulating Delta variant, across different clinical endpoints for different licensed vaccines under the context that obtaining such data from clinical trials and effectiveness studies may be difficult given the resources needed. We found that the predicted efficacy against the Delta variants are lower than that against prototype strains across different endpoints, with varying degrees depending on vaccine. Other variants of concern, such as the Beta variant, were also predicted to cause reductions in vaccine efficacy. Using data from studies of vaccine-induced neutralizing antibody responses, our study comprehensively predicted efficacy against multiple variants, potentially informing vaccine-related public-health decision-making.

We found that the predicted efficacy against the Delta variant suffered varying degrees of reduction across different clinical endpoints. The Delta variant harbors several key mutations of L452R, P681R, and T478K, which have been reported to be associated with immune evasion against viral neutralization.^12,13^ For example, the mutation L452R may promote interactions between the spike and angiotensin converting enzyme 2 (ACE2) receptor by inducing structural changes in binding domain,^14^ while the mutation T478K could cause the loss of neutralizing potency for some antibody lineages.^15^ We also found that the decline of predicted efficacy is more significant for adenovirus-vector vaccines than that of mRNA vaccines, likely due to higher overall immunogenicity of mRNA vaccines, and thus reductions of neutralizing antibodies against the Delta variant is less apparent.^8,16^

Comparing to other variants of concern, the loss of predicted efficacy of the Delta variant (relative to prototype strains) is less than that of the Beta variant, while their efficacy estimate is generally lower than the Alpha and Gamma variants across different clinical endpoints and vaccines (except for BNT162b2), similar to the variant-specific efficacy patterns reported by *Cromer et al*.^7^ Our findings include prediction of efficacies across additional vaccines, VOCs, and clinical endpoints, showing better protection with increasing levels of clinical severity.

Phase 3 randomized controlled trials provide accurate efficacy data among well-defined specific populations, while test-negative/cohort studies provide insights into protection in the real world among diverse populations, including those confronted with different SARS-CoV-2 variants. However, as most vaccine developers conducted prototype-virus-based clinical trials, and detected clinical outcomes by RT-PCR without performing sequencing, the efficacy or effectiveness of majority of vaccines against currently circulating SARS-CoV-2 variants are still largely unknown. In the absence of sequencing efforts in such studies, studies conducted in regions where a certain SARS-CoV-2 variant predominate may be able to approximate variant-specific protection, though accurate value would not be able to be ascertained without sequencing. Besides, most of the clinical trials were conducted before the Delta variant started taking over and now it is hard to do these studies again.

Given these circumstances, our study provides a timely and comprehensive estimation of predicted licensed-vaccine’s efficacy against WHO-designated VOCs based on neutralizing titers, which have been shown to be a highly predictive biomarker associated with vaccine protection.^9,17-19^ Based on the relationship between neutralizing antibodies and vaccine protection, using reduction-fold of titer from *in vitro* cross neutralizing assay to predict efficacy could be efficient and time-saving, and this framework could be easily extrapolated to other SARS-CoV-2 variants, which is undoubtedly conducive to outbreak preparedness by public health decision-makers.

Our study has several limitations. First, the predictions are based on the assumption that neutralizing antibodies are a primary determinant of immuno-protection, despite evidence that other immunologic mechanisms of humoral and cellular immunity may be important.^20^ However, previous studies showed that neutralizing antibody are highly predictive of immune protection from both symptomatic and asymptomatic infections.^9,18^ Second, we estimated the efficacy against infection by fitting a combined efficacy dataset of symptomatic and asymptomatic infection, due to the limited observed efficacy against infection alone, which may overestimate the predicted efficacy of preventing virus infection. Third, we used uncertainty intervals instead of true confidence intervals for the estimation due to provide a conservative and wider estimated interval. Finally, prediction of efficacy is based on a previously established model and it will be important to validate our results by conducting prospective studies in the real world.

In conclusion, our study predicts vaccine efficacy against different variants of concern across varied severity, focusing on the Delta variant. We confirm that existing vaccines based on prototype strains can provide protective efficacy against SARS-CoV-2 variants, especially for symptomatic and severe infections. We also provide the evidence that the Delta (and also Beta) variant are more likely to escape the immune protection induced by vaccines, warranting consideration of vaccines or therapeutics specific to those variants. Finally, as the various variants continue to emerge worldwide, our data may serve to inform decisions towards resource allocation and planning of mitigation measures by public health decision makers.

## Data Availability

All data used during the study are available from the corresponding author by request

## Contributors

H.Y. designed and supervised the study. X.C. did the literature search, set up the database and did all statistical analyses. X.C., A.S.A., and D.T.L. co-drafted the first version of the article. X.C., Z.C., R.S., W.L., N.Z., X.D., S.G., and helped with checking data and did the figures. D.T.L., A.S.A., J.Y, and H.Y. commented on the data and its interpretation, revised the content critically. All authors contributed to review and revision and approved the final manuscript as submitted and agree to be accountable for all aspects of the work.

## Acknowledgments

This study was funded by the Key Program of the National Natural Science Foundation of China (82130093) and National Science Fund for Distinguished Young Scholars (grant no. 81525023), the US National Institutes of Health (R01 AI135115 to D.T.L. and A.S.A.)

