## Supplements for "Prediction of vaccine efficacy of the Delta variant"

**Supplementary information**

### Methods

***Literature research and data extraction***

We conducted a systematic search from three peer-reviewed databases (PubMed, Web of Science and Embase) and an open science platform (Europe PMC), for studies published in English with predefined search terms **(Table S1)**. We included studies that are original analyses of COVID-19 vaccine efficacy with a randomized clinical trial design against wild type and variants, or are original analyses of COVID-19 vaccine effectiveness. Studies that were not in humans were excluded. We also excluded abstracts of congress meetings or conference proceedings, study protocols, media news, commentaries, and reviews. Detailed eligibility criteria conforming to the Population/Participants, Intervention, Comparator, Outcome, Setting/Study design (PICOS) format was summarized in **Table S2**. We screened all eligible studies to extract the following data: vaccine name, developer, study design, study location, study period, population size, age of participants, locally-circulating virus strains, overall and age-specific efficacy (full list of variables in **Tables S3 and S4)**. We previously reported a meta-analysis of *in vitro* neutralization titers of individuals who have been vaccinated with prototype-strain-based vaccines against both SARS-CoV-2 prototype strains and variants, and the data were continuously updated ^1^ We summarized geometric mean titers (GMTs) of neutralizing antibodies in vaccine recipients and the average reduction fold of GMTs in variants compared to the parental virus strains **(Table S5)**. The inclusion and exclusion of studies, screening and scrutinization of included studies, data extraction and verification were performed by two independent researchers, and a third researcher was consulted when disagreement arose.

**Data analysis**

For the collected efficacy data, only the efficacy after complete vaccination according to each vaccine’s immunization schedule (e.g., 7 days after two doses of BNT162b2/Pfizer, 14 days after two doses of mRNA-1273/Moderna) were used in analyses. For *in vitro* neutralizing antibody data, we prioritized using results from live virus neutralization assays, followed by those from pseudovirus neutralization assays. We also matched the doses and sampling time of subjects for which neutralizing antibody data were obtained and those in which efficacy data were obtained (e.g., BNT162b2/Pfizer vaccinees whose blood was taken within 7 days after the second dose are removed). For studies which aggregated sampling time data (e.g., using median and interquartile range to represent the sampling time for a group of people), we assumed all the people were sampled at the median/mean sampling time. For the estimation of age-specific efficacy, as age group categories were not exactly matched between efficacy data and neutralization data for mRNA-1273 vaccine, we assumed that the neutralization antibody levels for those older than 55 years old were similar to those older than 65 years.

**Statistical methods**

***Statistical model***

We followed the basic assumption that neutralizing antibodies is the best studied marker, and likely a major mechanism, of immune protection,^2^ though antiviral T and B cell memory likely contribute some degree of protection.^3,4^ Following the work by *Khoury et al*, *^2^* we predicted vaccine protection against variants with the following relationship between neutralizing antibody levels and vaccine efficacy **(equation 1)**, as well as an integral logistic-normal to calculate the probability of being protected **(equation 2)**.

$E_{I} \left( n | n_{50},k \right)=\frac{1}{1+e^{-k(n-n_{50})}}$*,* (1)

$P \left( n_{50},k, \mu_{s}, \sigma_{s} \right)=\int_{-\infty}^{+\infty} E_{I} \left( n | n_{50},k \right) f(n|\mu_{s},\sigma_{s})dn,$(2)

A logistic model was assumed to model the relationship for equation (1), where *E_I_* is the vaccine efficacy given the log-transformed neutralizing antibody titer *n*. *n_50_* is the neutralization titer at which an individual will have a 50% protective efficacy. The parameter *k* controls the steepness of the logistic function.^2^ For equation (2), assuming that neutralizing antibodies follow a normal distribution with mean *μ_s_* and standard deviation *σ_s_* (here the pooled standard deviation estimated from a combined dataset of neutralization titers from all studies was used)^2^, the *f* indicates the probability density function of neutralization titer. $P(\cdot)$ represents the proportion of vaccinated population for a study *s* that will be protected.^2^

*Estimating efficacy of variants against COVID-19 symptomatic and severe case*

Using previous **equations (1)**, we firstly modelled the relationship between efficacy against the prototype strain and neutralizing titers as *Khoury et al*., described.^2^ Then, distributions of fold changes in neutralizing antibodies for different variants were added into the model to predict changes in efficacy for each vaccine. For the parameters used in this model, the slope *k* for COVID-19 symptomatic and severe cases, and pooled standard deviation *σ_s_* were the same as reported by *Khoury et al*.

*Estimating efficacy of variants against SARS-CoV-2 infection*

For the estimation of efficacy against infection, firstly, we used the binomial probability function and likelihood **(equation 3)** to estimate the parameter *k and n_50_* by fitting the efficacy data of infections and symptomatic cases from phase 3 clinical trials (due to limited number of studies for infection alone) according to methods reported by *Khoury et al*.^2^

$L_{s}\left( N_{s}^{c},I_{s}^{c},N_{s}^{v},I_{s}^{v}{,\mu}_{s}, \sigma_{s} \right|n_{50},k,b_{s})=Bi(N_{s}^{c},I_{s}^{c},b_{s})Bi(N_{s}^{v},I_{s}^{v},b_{s}(1-P(n_{50},k, \mu_{s}, \sigma_{s})))$*,* (3)

where *b_s_* is the probability of an unvaccinated individual (control) being infected

in study *s*, *b_s_(1−P(n_50_, k, μ_s_, σ_s_))* is the probability of infection in

the vaccinated individuals and *Bi(N, K, p)* is the binomial probability

mass function of the probability of *K* events from a sample size of *N*, for which

each event has a probability *P*. Second, similar to estimation of efficacy against symptomatic and severe, the fold-change of neutralizing antibody level of variants were added to predict efficacy against SARS-CoV-2 infection according to both **equation (1)** and **equation (2)**.

*Estimating uncertainty intervals*

We used the 95% confidence intervals of fold-change of neutralizing titers, combined with the 95% confidence interval of efficacy against prototype strains, and selected the highest and the lowest estimates to be the upper and lower limits of our uncertainty intervals. All statistical analyses were done using R (version 4.0.1).

### Supplementary Tables

#### Table S1. Search strategy for studies that reported COVID-19 vaccine efficacy and effectiveness

| Database | Step | Search strategy | Number of articles* |
| --- | --- | --- | --- |
| Pubmed | #1 | COVID-19[All Fields] | 143,932 |
|  | #2 | vaccin*[Title/Abstract] or immunization[Title/Abstract] | 387,536 |
|  | #3 | #1 AND #2 | 11,504 |
|  | #4 | COVID-19 Vaccines[MeSH Terms] | 2,954 |
|  | #5 | #3 OR #4 | 12,046 |
|  | #6 | BNT162b2[All Fields] OR Pfizer-BioNTech[All Fields] OR Pfizer/BioNTech[All Fields] OR Comirnaty[All Fields] | 480 |
|  | #7 | mRNA-1273[All Fields] OR Moderna COVID-19 vaccine[All Fields] | 179 |
|  | #8 | ChAdOx1 nCoV-19[All Fields] OR AZD1222[All Fields] OR ChAdOx1-S[All Fields] OR Oxford-AstraZeneca[All Fields] OR Covishield[All Fields] | 186 |
|  | #9 | Gam-COVID-Vac[All Fields] OR Sputnik V[All Fields] | 25 |
|  | #10 | Ad5 nCoV[All Fields] OR Cansino[All Fields] | 130 |
|  | #11 | BBIBP-CorV[All Fields] OR WBIP[All Fields] OR Sinopharm[All Fields] | 145 |
|  | #12 | CoronaVac[All Fields] OR SinoVac[All Fields] | 105 |
|  | #13 | BBV152[All Fields] OR Covaxin[All Fields] OR Bharat Biotech[All Fields] | 69 |
|  | #14 | Ad26.COV2.S[All Fields] OR Janssen COVID-19 Vaccine[All Fields] | 49 |
|  | #15 | ZF2001[All Fields] OR Anhui Zhifei Longcom[All Fields] | 9 |
|  | #16 | EpiVacCorona[All Fields] | 1 |
|  | #17 | KCONVAC[All Fields] | 1 |
|  | #18 | #6 OR #7 OR #8 OR #9 OR #10 OR #11 OR #12 OR #13 OR #14 OR #15 OR #16 OR #17 | 1,149 |
|  | #19 | #5 OR #18 | 12,450 |
|  | #20 | Immunogenicity, Vaccine[MeSH Terms] | 2,065 |
|  | #21 | Immunity[MeSH Terms] | 395,451 |
|  | #22 | immunogenic*[Title/Abstract] | 65,142 |
|  | #23 | effic*[Title/Abstract] | 1,924,784 |
|  | #24 | effect*[Title/Abstract] | 7,388,003 |
|  | #25 | immune*[Title/Abstract] | 692,578 |
|  | #26 | respons*[Title/Abstract] | 3,396,441 |
|  | #27 | protect*[Title/Abstract] | 864,520 |
|  | #28 | #20 OR #21 OR #22 OR #23 OR #24 OR #25 OR #26 OR #27 | 10,982,066 |
|  | #29 | #19 AND #28 | 7,825 |
| Web of Science | #1 | TS = COVID-19 | 172,501 |
|  | #2 | TS = vaccin* | 658,077 |
|  | #3 | #1 AND #2 | 13,138 |
|  | #4 | TS = (BNT162b2 OR Pfizer-BioNTech OR Pfizer/BioNTech OR Comirnaty) | 456 |
|  | #5 | TS = (mRNA-1273 OR Moderna COVID-19 vaccine) | 287 |
|  | #6 | TS = (ChAdOx1 nCoV-19 OR AZD1222 OR ChAdOx1-S OR Oxford-AstraZeneca OR Covishield) | 172 |
|  | #7 | TS = (Gam-COVID-Vac OR Sputnik V) | 71 |
|  | #8 | TS = (Ad5 nCoV OR Cansino) | 37 |
|  | #9 | TS = (BBIBP-CorV OR WBIP OR Sinopharm) | 35 |
|  | #10 | TS = (CoronaVac OR SinoVac) | 58 |
|  | #11 | TS = (BBV152 OR Covaxin OR Bharat Biotech) | 44 |
|  | #12 | TS = (Ad26.COV2.S OR Janssen COVID-19 Vaccine) | 62 |
|  | #13 | TS = (ZF2001 OR Anhui Zhifei Longcom) | 6 |
|  | #14 | TS = (EpiVacCorona) | 3 |
|  | #15 | TS = (KCONVAC) | 1 |
|  | #16 | #4 OR #5 OR #6 OR #7 OR #8 OR #9 OR #10 OR #11 OR #12 OR #13 OR #14 OR #15 | 945 |
|  | #17 | #3 OR #16 | 13,378 |
|  | #18 | TS = immunogenic* | 122,444 |
|  | #19 | TS = effic* | 12,789,042 |
|  | #20 | TS = effect* | 31,459,770 |
|  | #21 | TS = immune* | 3,671,086 |
|  | #22 | TS = repons* | 11,044 |
|  | #23 | TS = protect* | 5,459,212 |
|  | #24 | #18 OR #19 OR #20 OR #21 OR #22 OR #23 | 45,061,106 |
|  | #25 | #17 AND #24 | 8,553 |
| Embase | #1 | coronavirus disease 2019'/de | 123,978 |
|  | #2 | COVID-19 | 127,066 |
|  | #3 | #1 OR #2 | 143,296 |
|  | #4 | vaccin* | 561,441 |
|  | #5 | #3 AND #4 | 12,442 |
|  | #6 | bnt162b2 OR 'Pfizer BioNTech' OR Comirnaty | 395 |
|  | #7 | mRNA-1273 OR 'Moderna COVID-19 vaccine' | 267 |
|  | #8 | ChAdOx1 nCoV-19' OR AZD1222 OR ChAdOx1-S OR 'Oxford AstraZeneca' OR Covishield | 275 |
|  | #9 | Gam-COVID-Vac OR 'Sputnik V' | 85 |
|  | #10 | Ad5 nCoV' OR Cansino | 397 |
|  | #11 | BBIBP-CorV OR WBIP OR Sinopharm | 833 |
|  | #12 | CoronaVac OR SinoVac | 311 |
|  | #13 | BBV152 OR Covaxin OR 'Bharat Biotech' | 202 |
|  | #14 | Ad26.COV2.S OR 'Janssen COVID-19 Vaccine' | 73 |
|  | #15 | ZF2001 OR 'Anhui Zhifei Longcom' | 26 |
|  | #16 | EpiVacCorona | 10 |
|  | #17 | KCONVAC | 1 |
|  | #18 | #6 OR #7 OR #8 OR #9 OR #10 OR #11 OR #12 OR #13 OR #14 OR #15 OR #16 OR #17 | 2,186 |
|  | #19 | #5 OR #18 | 13,857 |
|  | #20 | immunogenic* | 107,428 |
|  | #21 | effic* | 3,077,373 |
|  | #22 | effect* | 10,582,126 |
|  | #23 | immune* | 1,251,886 |
|  | #24 | repons* | 8,581 |
|  | #25 | protect* | 1,363,644 |
|  | #26 | #20 OR #21 OR #22 OR #23 OR #24 OR #25 | 13,288,803 |
|  | #27 | #19 AND #26 | 8,581 |
| Europe PMC | #1 | COVID-19 AND vaccin* | 54,038 |
|  | #2 | immunogenic* OR effect* OR effic* OR immune* OR protect* OR protect* | 13,826,389 |
|  | #3 | Type: Preprints | 307,380 |
|  | #4 | #1 AND #2 AND #3 | 11,583 |

#### Table S2. Inclusion and exclusion criteria of searched studies

| Population or participants | Population without immunosuppressive conditions, including females and/or males of any age group, with or without prior SARS-CoV-2 infection, exclude HIV-positive, cancer, Organ Transplant Recipients, Pregnant Women and so on. |
| --- | --- |
| Intervention or exposure | COVID-19 candidate vaccine (BNT162b2, mRNA-1273, BBIBP-CorV, WBIP, CoronaVac, BBV152, ChAdOx1 nCoV-19, Gam-COVID-Vac, Ad5 nCoV and so on) |
| Comparison groups | 1) Placebo, and/or other vaccine adminstrate  2) No COVID-19 vaccination. |
| Primary outcomes | Efficacy and Effectiveness against SARS-CoV-2 (wild type and variants) associated clinical outcomes (including, but not limited to, symptomatic case, medical attendance, death) |
| Setting | Any setting within any geographical location. |
| Study designs | Any study including participants who received COVID-19 vaccine through a clinical trial, and where data were presented by number of doses received. This could include the original clinical trial in which vaccine was administered or subsequent observational studies, e.g. prospective cohort, cross-sectional studies or case-control studies. |
| Language | Restricted to English-language publications. |
| Exclusion criteria | Studies not meeting inclusion criteria were excluded. Additionally:   1) Studies that were not in humans. 2) review, modelling study |

#### Table S3. Summary of vaccine efficacy from included studies

| **Vaccine** | **Developer** | **Study location** | **Population size** | **Age range** | **Design/Measure of effect** | **Circulation of VOCs** | **Period** | **Against infection** | **Against symptomatic case** | **Against hospitalization** | **Against severe outcome** |
| --- | --- | --- | --- | --- | --- | --- | --- | --- | --- | --- | --- |
| **mRNA** | | | | | | | | | | | |
| BNT162b2 | BioNTech/Pfizer | United States^5^ | 2,260 | 12-15 years | RCT/Efficacy | Prototype | 7 days after dose 2 | - | 100.0 (75.3-100) | - | - |
|  |  | United States, Argentina, Brazil, South Africa^6^ | 43,355 | ≥16 years |  |  | After dose 1* | - | 82.0 (75.6-86.9) | - | 88.9 (20.1-99.7) |
|  |  |  | 37,706 | ≥16 years |  |  | 7 days after dose 2 | - | 95.0 (90.3-97.6) | - | 75.0 (-152.6, 99.5) |
|  |  |  | 21,785 | 16-55 years |  |  |  | - | 95.6 (89.4-98.6) | - | - |
|  |  |  | 15,921 | > 55 years |  |  |  | - | 94.7 (66.7-99.9) | - | - |
|  |  |  |  |  |  |  |  | - |  | - | - |
|  |  |  |  |  |  |  |  | - |  | - | - |
|  |  | United States, Argentina, Brazil, South Africa^7^ |  | ≥ 12 years | RCT/Efficacy | Mixed | 7 days after dose 2 | - | 91.2 (88.9, 93.0) | - | 95.7 (73.9-99.9) |
|  |  |  |  | 16-17 years |  |  |  | - | 100.0 (58.0-100.0) | - | - |
|  |  |  |  | 16-55 years |  |  |  | - | 91.2 (88.3-93.5) | - | - |
|  |  |  |  | > 55 years |  |  |  | - | 90.9 (86.3-94.2) | - | - |
|  |  |  |  | ≥ 65 years |  |  |  | - | 94.5 (88.3-97.8) | - | - |
|  |  |  |  | ≥ 75 years |  |  |  | - | 96.2 (76.9-99.9) | - | - |
| mRNA-1273 | Moderna | United States^8^ | 3,700 | 12-17 years | RCT/Efficacy | Prototype | 14 days after dose 2 | - | 100.0 | - | - |
|  |  |  | 28,207 | ≥18 years |  |  |  | 89.6 (85.2-92.6)* | 94.1 (89.3-96.8) | - | 100.0 (NE-100.0) |
|  |  |  | 21,072 | 18-65 years |  |  |  | - | 95.6 (90.6-97.9) | - | - |
|  |  |  | 7,135 | ≥65 years |  |  |  | - | 86.4 (61.4-95.2) | - | - |
| **Inactivated** | | | | | | | | | | | |
| CoronaVac | SinoVac | Brazil^9^ | 12,688 | >18 years | RCT/Efficacy | Prototype, P.1 | 14 days after dose 2 | - | 50.7 (35.9-62.0) | 100 (56-100) | 100.0 (16.9-100.0) |
|  |  |  | 9,404 | 18-59 years |  |  |  | - | 50.7 (35.8-62.1) | - | - |
|  |  |  | 419 | ≥ 60 years |  |  |  | - | 51.1 (-166.9-91.0) | - |  |
|  |  | Brazil^10^ |  | >18 years |  | B.1.1.28 |  | - | 73 (46-86) | - | - |
|  |  |  |  | >18 years |  | P.2 |  | - | 69 (55-78) | - | - |
|  |  |  |  | >18 years |  | B.1.1.33 |  | - | 88.2 (5-99) | - | - |
|  |  |  |  | >18 years |  | P.1 |  | - | 64 (-2-87) | - | - |
|  |  | Turkey^11^ | 10,218 | ≥18 years |  | Prototype |  | - | 83.5 (65.4-92.1) | - | 100 (20.4-100) |
|  |  | Indonesia^12^ | 1,602 | ≥18 years |  |  |  | - | 65 (20-85) | - | - |
| BBIBP-Corv | Sinopharm | Multi-country^13^ | 12,726 | ≥18 years | RCT/Efficacy | Prototype | 14 days after dose 2 | 73.5 (60.6-82.2) | 78.1 (64.8-86.3) | - | 100 |
|  |  |  | 12,525 | 18-59 years |  |  |  |  | 78.1 (64.9-86.4) | - | - |
| WBIP | Sinopharm | Multi-country^13^ | 12,734 | ≥18 years | RCT/Efficacy | Prototype | 14 days after dose 2 | 64.0 (48.8-74.7) | 72.8 (58.1-82.4) | - | 100 (NE) |
|  |  |  | 12,530 | 18-59 years |  |  |  | - | 72.8 (58.0-82.4) | - | - |
| BBV152 | Bharat Biotech | India^14^ | 16,973 | ≥18 years | RCT/Efficacy | Mixed | 14 days after dose 2 | 68.8 (46.7-82.5) | 77.8 (65.2-86.4) | - | 93.4 (57.1-99.8) |
|  |  |  | 15,115 | 18-59 years |  | Mixed |  | - | 79.4 (66.0-88.2) | - | - |
|  |  |  | 1,858 | ≥60 years |  | Mixed |  | - | 67.8 (8.0-90.0) | - | - |
|  |  |  | 16,973 | ≥18 years |  | B.1.617.2 (Delta) |  | - | 65.2 (33.1-83.0) | - | - |
|  |  |  | 16,973 | ≥18 years |  | B.1.617.1 (Kappa) |  | - | 90.1 (30.4-99.8) | - | - |
| **Non-replicating vector** | | | | | | | | | | | |
| Gam-COVID-Vac | Russian Gamaleya NRCEM | Russia^15^ | 18,695 | ≥18 years | RCT/Efficacy | Prototype | 14 days after dose 2 | - | 91.1 (83.8-95.1) | - | 100.0 (94.4-100.0) |
| Ad26.COV2.S | Janssen | United States, Brazil, South Africa^16^ | 39,058 | ≥18 years | RCT/Efficacy | Prototype, P.1, B.1.351 | 14 days after dose 1 | - | 66.9 (59.1-73.4) | - | 76.7 (54.6-89.1) |
|  |  |  | 29,111 | 18-59 years |  |  |  | - | 65.8 (56.2-73.1) | - | - |
|  |  |  | 14,672 | ≥ 60 years |  |  |  | - | 74.5 (57.9-84.3) | - | - |
|  |  | South Africa | 4,969 | ≥18 years | RCT/Efficacy | B.1.351 | 14 days after dose 1 | - | 52.0 (30.3-67.4) | - | 73.1 (40.0-89.4) |
|  |  | United States, Brazil, South Africa^16^ | 39,058 | ≥18 years | RCT/Efficacy | Prototype, P.1, B.1.351 | 28 days after dose 1 | - | 66.1 (55.0-74.8) | - | 85.4 (54.2-96.9) |
|  |  |  | 29,111 | 18-59 years |  |  |  | - | 69.3 (57.4-77.7) | - | - |
|  |  |  | 14,672 | ≥ 60 years |  |  |  | - | 67.9 (38.2-82.8) | - | - |
|  |  | South Africa^16^ | 4,969 | ≥18 years | RCT/Efficacy | B.1.351 | 28 days after dose 1 | - | 64.0 (41.2-78.7) | - | 81.7 (46.2-95.4) |
| ChAdOx1-S | AstraZeneca | United Kingdom, Brazil, South Africa^17^ | 12,604 | ≥18 years | RCT/Efficacy | Prototype | 21 days after dose 1 | 46.3 (31.8-57.8) | 58.3 (44.0-68.9) | 100 | 100 |
|  |  |  | 11,636 | ≥18 years |  |  | 14 days after dose 2 | 55.7 (41.1-66.7) | 70.4 (54.8-80.6) | 100 | 100 |
|  |  | South Africa^18^ | 1,464 | ≥18 years | RCT/Efficacy | B.1.351 | 14 days after dose 2 | - | 10.4 (-76.8-54.8) | - | - |
|  |  | United Kingdom^19^ | 8,534 | ≥18 years | RCT/Efficacy | B.1.1.7 | 14 days after dose 2 | 61.7 (36.7-76.9) | 70.4 (43.6-84.5) | 100 | - |
| **Protein subunit** | | | | | | | | | | | |
| NVX-CoV2373 | Novavax | United Kingdom^20^ | 14,039 | ≥18 years | RCT/Efficacy | Non-B.1.1.7 | 7 days after dose 2 | - | 96.4 (73.6-99.5) | 100 | 100 |
|  |  |  | 14,039 | ≥18 years |  | B.1.1.7 |  | - | 86.3 (71.3-93.5) | 100 | 100 |
|  |  | South Africa^21^ | 2,536 | ≥18 years |  | B.1.351 |  | - | 60.1 (19.9-80.1) | - | - |

#### Table S4. Summary of vaccine effectiveness from included studies

| **Vaccine** | **Developer** | **Study location** | **Population size** | **Age range** | | **Design/Measure of effect** | | **Circulation of VOCs** | | **Period** | | **Against infection** | | **Against symptomatic case** | | **Against hospitalization** | | **Against severe outcome** |
| --- | --- | --- | --- | --- | --- | --- | --- | --- | --- | --- | --- | --- | --- | --- | --- | --- | --- | --- |
| **mRNA** | | | | | | | | | | | | | | | | | | |
| BNT162b2 | BioNTech/Pfizer | United Kingdom^22,23^ | 11,621 | ≥16 years | | test-negative/Effectiveness | | B.1.1.7 | | 21 days after dose 1 | | - | | 47.5 (41.6-52.8) | | 83 (62-93) | | - |
|  |  |  | 11,621 | ≥16 years | |  |  | B.1.1.7 | | 14 days after dose 2 | | - | | 93.7 (91.6-95.3) | | 95 (78-99) | | - |
|  |  |  | 1,054 | >16 years | |  |  | B.1.1.7 | | 21 days after dose 1 | | - | | 35.6 (22.7-46.4) | | 94 (46-99) | | - |
|  |  |  | 1,054 | >16 years | |  |  | B.1.1.7 | | 14 days after dose 2 | | - | | 88.0 (85.3-90.1) | | 96 (86-99) | | - |
|  |  |  |  | 18-64 years | |  |  | B.1.617.2 | | 14 days after dose 2 | | 82 (79-85) | | 86 (83-88) | | - | | - |
|  |  | Qatar^24^ | 37,934 | ≥16 years | |  |  |  | | After dose 1 | | 29.5 (22.9-35.5) | | - | | - | | 54.1 (26.1-71.9) |
|  |  |  | 32,808 | ≥16 years | |  |  |  | | 14 days after dose 2 | | 89.5 (85.9-92.3) | | - | | - | | 100.0 (81.7-100.0) |
|  |  |  | 43,012 | ≥16 years | |  |  | B.1.351 | | After dose 1 | | 16.9 (10.4-23.0) | | - | | - | | 0.0 (0.0-19.0) |
|  |  |  | 39,150 | ≥16 years | |  |  | B.1.351 | | 14 days after dose 2 | | 75.0 (70.5-78.9) | | - | | - | | 100.0 (73.7-100.0) |
|  |  |  |  | ≥16 years | |  |  | B.1.617.2 | | After dose 1 | | 65.5 (40.9-79.9) | | 76.3 (46.7-90.7) | | - | | 100 (NE) |
|  |  |  |  | ≥16 years | |  |  | B.1.617.2 | | 14 days after dose 2 | | 59.6 (50.7-66.9) | | 56.1 (41.4-67.2) | | - | | 97.3 (84.4-95.5) |
|  |  | Scotland^25^ | 199,375 | ≥16 years | | test-negative/Effectiveness | | B.1.617.2 | | 0-27 days after dose 1 | | 12 | | 18 | | - | | - |
|  |  |  | 199,375 | ≥16 years | |  |  |  |  | 28 days after dose 1 | | 30 | | 33 | | - | | - |
|  |  |  | 199,375 | ≥16 years | |  |  |  |  | 0-13 days after dose 2 | | 66 | | 84 | | - | | - |
|  |  |  | 199,375 | ≥16 years | |  |  |  |  | 14 days after dose 2 | | 79 | | 83 | | - | | - |
|  |  |  | 199,375 | ≥16 years | |  |  | B.1.1.7 | | 0-27 days after dose 1 | | 31 | | 28 | | - | | - |
|  |  |  | 199,375 | ≥16 years | |  |  |  |  | 28 days after dose 1 | | 38 | | 27 | | - | | - |
|  |  |  | 199,375 | ≥16 years | |  |  |  |  | 0-13 days after dose 2 | | 73 | | 78 | | - | | - |
|  |  |  | 199,375 | ≥16 years | |  |  |  |  | 14 days after dose 2 | | 92 | | 92 | | - | | - |
|  |  | Canada^23^ |  | ≥16 years | | test-negative/Effectiveness | | B.1.1.7 | | 14 days after dose 1 | | - | | 66 (64-68) | | 80 (78-82) | | - |
|  |  |  |  | ≥16 years | |  |  |  |  | 21 days after dose 1 | | - | | 69 (67-71) | | 85 (83-86) | | - |
|  |  |  |  | ≥16 years | |  |  |  |  | 7 days after dose 2 | | - | | 89 (86-91) | | 95 (9-97) | | - |
|  |  |  |  | ≥16 years | |  |  |  |  | 14 days after dose 2 | | - | | 89 (87-91) | | 96 (93-98) | | - |
|  |  |  |  | ≥16 years | |  |  | B.1.351/P.1 | | 14 days after dose 1 | | - | | 60 (52-67) | | 77 (69-83) | | - |
|  |  |  |  | ≥16 years | |  |  |  |  | 21 days after dose 1 | | - | | 65 (56-71) | | 83 (75-88) | | - |
|  |  |  |  | ≥16 years | |  |  |  |  | 7 days after dose 2 | | - | | 84 (69-92) | | 95 (81-99) | | - |
|  |  |  |  | ≥16 years | |  |  |  |  | 14 days after dose 2 | | - | | 85 (70-93) | | 98 (82-100) | | - |
|  |  |  |  | ≥16 years | |  |  | B.1.617.2 | | 14 days after dose 1 | | - | | 56 (45-64) | | 78 (65-86) | | - |
|  |  |  |  | ≥16 years | |  |  |  |  | 21 days after dose 1 | | - | | 61 (51-70) | | 78 (64-87) | | - |
|  |  |  |  | ≥16 years | |  |  |  |  | 7 days after dose 2 | | - | | 87 (64-95) | | - | | - |
|  |  |  |  | ≥16 years | |  |  |  |  | 14 days after dose 2 | | - | | 85 (59-94) | | - | | - |
| mRNA-1273 | Moderna | Qatar^26^ |  | ≥18 years | | test-negative/Effectiveness | | B.1.1.7 | | 14 days after dose 1 | | 88.1 (83.7-91.5) | | - | | - | | - |
|  |  |  |  | ≥18 years | |  |  |  |  | 14 days after dose 2 | | 100.0 (91.8-100) | | - | | - | | - |
|  |  |  |  | ≥18 years | |  |  | B.1.351 | | 14 days after dose 1 | | 61.3 (56.5-65.5) | | - | | - | | - |
|  |  |  |  | ≥18 years | |  |  |  |  | 14 days after dose 2 | | 96.4 (91.9-98.7) | | - | | - | | - |
|  |  | Canada^27^ |  | ≥18 years | |  |  | B.1.1.7 | | 14 days after dose 1 | | - | | - | | - | | - |
|  |  |  |  | ≥18 years | |  |  |  |  | 21 days after dose 1 | | - | | - | | - | | - |
|  |  |  |  | ≥18 years | |  |  |  |  | 7 days after dose 2 | | - | | - | | - | | - |
|  |  |  |  | ≥18 years | |  |  |  |  | 14 days after dose 2 | | - | | - | | - | | - |
|  |  |  |  | ≥18 years | |  |  | B.1.351/P.1 | | 14 days after dose 1 | | - | | - | | - | | - |
|  |  |  |  | ≥18 years | |  |  |  |  | 21 days after dose 1 | | - | | - | | - | | - |
|  |  |  |  | ≥18 years | |  |  | B.1.617.2 | | 14 days after dose 1 | | - | | - | | - | | - |
|  |  |  |  | ≥18 years | |  |  |  |  | 21 days after dose 1 | | - | | - | | - | | - |
| **Inactivated** | | | | | | | | | | | | | | | |  | |  |
| CoronaVac | SinoVac | Brazil^28^ | 2,797 | ≥18 years | | test-negative/Effectiveness | | P.1 | | 14 days after dose 1 | | 35.1 (-6.6-60.5) | | 49.6 (11.3-71.4) | | - | | - |
|  |  |  | 1,665 | ≥18 years | |  |  |  |  | 14 days after dose 2 | | 37.9 (-46.4-73.6) | | 36.8 (-54.9-74.2) | | - | | - |
|  |  | Brazil^29^ |  | ≥70 years | | test-negative/Effectiveness | | P.1 | | 14 days after dose 2 | | - | | 41.6 (26.9-53.3) | | 59.0 (44.2-69.8) | | 71.4 (53.7-82.3) (death) |
|  |  |  |  | 70-74 years | |  |  |  |  |  |  | - | | 61.8 (34.8-77.7) | | 80.1 (55.7-91.0) | | 86.0 (50.4-96.1) (death) |
|  |  |  |  | 75-79 years | |  |  |  |  |  |  | - | | 48.9 (23.3-66.0) | | 69.5 (42.4-83.8) | | 87.1 (60.2-95.8) (death) |
|  |  |  |  | ≥80 years | |  |  |  |  |  |  | - | | 28.0 (0.60-47.9) | | 43.4 (15.4-62.0) | | 49.9 (8.1-72.7) (death) |
| **Non-replicating vector** | | | | | | | | | | | | | | |  | |  | |
| ChAdOx1-S | AstraZeneca | United Kingdom^22,23^ | 11,621 | ≥18 years | test-negative/Effectiveness | | B.1.1.7 | | 21 days after dose 1 | | - | | 48.7 (45.2-51.9) | | 76 (61-85) | | - | |
|  |  |  | 11,621 | ≥18 years |  |  |  |  | 14 days after dose 2 | | - | | 74.5 (68.4-79.4) | | 86 (53-96) | | - | |
|  |  |  | 1,054 | ≥18 years |  |  | B.1.617.2 | | 21 days after dose 1 | | - | | 30.0 (24.3-35.3) | | 71 (51-83) | | - | |
|  |  |  | 1,054 | ≥18 years |  |  |  |  | 14 days after dose 2 | | - | | 67.0 (61.3-71.8) | | 92 (75-97) | | - | |
|  |  | Scotland^25^ | 199,375 | ≥18 years | test-negative/Effectiveness | | B.1.617.2 | | 0-27 days after dose 1 | | 7 | | 23 | | - | | - | |
|  |  |  | 199,375 | ≥18 years |  |  |  |  | 28 days after dose 1 | | 18 | | 33 | | - | | - | |
|  |  |  | 199,375 | ≥18 years |  |  |  |  | 0-13 days after dose 2 | | 25 | | 37 | | - | | - | |
|  |  |  | 199,375 | ≥18 years |  |  |  |  | 14 days after dose 2 | | 60 | | 61 | | - | | - | |
|  |  |  | 199,375 | ≥18 years |  |  | B.1.1.7 | | 0-27 days after dose 1 | | 9 | | 17 | | - | | - | |
|  |  |  | 199,375 | ≥18 years |  |  |  |  | 28 days after dose 1 | | 37 | | 39 | | - | | - | |
|  |  |  | 199,375 | ≥18 years |  |  |  |  | 0-13 days after dose 2 | | 64 | | 65 | | - | | - | |
|  |  |  | 199,375 | ≥18 years |  |  |  |  | 14 days after dose 2 | | 73 | | 81 | | - | | - | |
|  |  | Brazil^30^ |  | ≥60 years | test-negative/Effectiveness | | P.1 | | 28 days after dose 1 | | - | | 33.4 (26.4-39.7) | | 55.1 (46.6-62.2) | | ICU admission: 50.9 (-41.8-83) Invasive mechanical ventilation: 70.5 (54.9-80.8) Death: 61.8 (48.9-71.4) | |
|  |  |  |  | ≥60 years |  |  |  |  | 14 days after dose 2 | | - | | 77.9 (69.2-84.2) | | 87.6 (78.2-92.9) | | ICU admission: 89.9 (70.9-96.5) Invasive mechanical ventilation: 96.5 (81.7-99.3) Death: 93.6 (81.9-97.7) | |
|  |  | Canada^27^ |  | ≥18 years | test-negative/Effectiveness | | B.1.1.7 | | 14 days after dose 1 | | - | | 64 (60-68) | | 85 (81-88) | | - | |
|  |  |  |  | ≥18 years |  | |  |  | 21 days after dose 1 | | - | | 72 (68-76) | | 90 (86-93) | | - | |
|  |  |  |  | ≥18 years |  | |  |  | 7 days after dose 2 | | - | | 79 (-57-97) | | 67 (-155-96) | | - | |
|  |  |  |  | ≥18 years |  | |  |  | 14 days after dose 2 | | - | | 75 (-98-97) | | - | | - | |
|  |  |  |  | ≥18 years |  | | B.1.351/P.1 | | 7 days after dose 2 | | - | | 48 (28-63) | | 83 (66-92) | | - | |
|  |  |  |  | ≥18 years |  | |  |  | 14 days after dose 2 | | - | | 50 (27-66) | | 82 (91-92) | | - | |
|  |  |  |  | ≥18 years |  | | B.1.617.2 | | 7 days after dose 2 | | - | | 67 (44-80) | | 88 (60-96) | | - | |
|  |  |  |  | ≥18 years |  | |  |  | 14 days after dose 2 | | - | | 70 (46-83) | | 87 (56-96) | | - | |

#### Table S5. Fold change of neutralization antibody level used in this study^1^

| Vaccine | Variants (Pango lineage) | Fold change of neutralization titer  (95%CI) | Type of neutralization assay |
| --- | --- | --- | --- |
| BNT162b2 | Alpha (B.1.1.7) | 1.13 (1.02-1.25) | Live virus neutralization assay |
| BNT162b2 | Beta (B.1.351) | 4.63 (4.20-5.10) | Live virus neutralization assay |
| BNT162b2 | Gamma (P.1) | 1.98 (1.65-2.36) | Live virus neutralization assay |
| BNT162b2 | Delta (B.1.617.2) | 0.99 (0.81-1.20) | Live virus neutralization assay |
| mRNA-1273 | Alpha (B.1.1.7) | 0.87 (0.61-1.25) | Live virus neutralization assay |
| mRNA-1273 | Beta (B.1.351) | 5.33 (4.07-6.98) | Live virus neutralization assay |
| mRNA-1273 | Delta (B.1.617.2) | 3.37 (2.21-5.15) | Live virus neutralization assay |
| Ad26.COV2.S | Alpha (B.1.1.7) | 0.61 (0.41-0.93) | Lentivirus-vector pseudovirus neutralization assay |
| Ad26.COV2.S | Beta (B.1.351) | 3.05 (2.16-4.31) | Lentivirus-vector pseudovirus neutralization assay |
| Ad26.COV2.S | Gamma (P.1) | 1.30 (0.74-2.26) | Lentivirus-vector pseudovirus neutralization assay |
| Ad26.COV2.S | Delta (B.1.617.2) | 2.54 (1.84-3.49) | Lentivirus-vector pseudovirus neutralization assay |
| ChAdOx1 nCoV-19 | Alpha (B.1.1.7) | 0.74 (0.54-1.02) | Live virus neutralization assay |
| ChAdOx1 nCoV-19 | Beta (B.1.351) | 3.55 (2.50-5.04) | Live virus neutralization assay |
| ChAdOx1 nCoV-19 | Gamma (P.1) | 1.38 (0.96-1.99) | Live virus neutralization assay |
| ChAdOx1 nCoV-19 | Delta (B.1.617.2) | 3.49 (2.44-4.97) | Live virus neutralization assay |
| Gam-COVID-Vac | Alpha (B.1.1.7) | 0.60 (0.34-1.06) | VSV-vector pseudovirus neutralization assay |
| Gam-COVID-Vac | Beta (B.1.351) | 4.78 (2.04-11.22) | VSV-vector pseudovirus neutralization assay |
| NVX-CoV2373 | Alpha (B.1.1.7) | 2.08 (1.18-3.66) | Lentivirus-vector pseudovirus neutralization assay |
| NVX-CoV2373 | Beta (B.1.351) | 18.50 (11.47-29.86) | Lentivirus-vector pseudovirus neutralization assay |

#### Table S6. Model parameters used in prediction of efficacy

| Model structure | LL | AIC | Slope (***k***) | Pooled SD ($\sigma_{s}$)^a^ |
| --- | --- | --- | --- | --- |
| Fitting protection from symptomatic vs severe COVID-19 | | | | |
| Different 𝒏_𝟓𝟎_ ^a^ | -66.08 | 138.16 | 2.94 (2.14-4.04) | 0.44 |
| Fitting protection from symptomatic COVID-19 vs SARS-CoV-2 infection | | | | |
| Different 𝒏_𝟓𝟎_ | -61.10 | 128.20 | 2.88 (2.19-3.78) | 0.44 |
| Same 𝒏_𝟓𝟎_ | -70.09 | 144.18 | 2.31 (1.80-2.96) | 0.44 |

a indicates the parameter was extracted from *Khoury et al.^2^*

#### Table S7. Comparison of predicted efficacy and efficacy/effectiveness in published data.

| Vaccine | Variants  (Pango lineage) | Efficacy in published data (%) | Effectiveness in published data (%) | Predicted efficacy (%) |
| --- | --- | --- | --- | --- |
| Against SARS-CoV-2 infection | | | | |
| ChAdOx1 nCoV-19 | Alpha (B.1.1.7) | United Kingdom: 61.7 (36.7-76.9) | Scotland: 73 | 62.7 (40.7-78.7) |
| ChAdOx1 nCoV-19 | Delta (B.1.617.2) | - | Scotland: 60  United Kingdom: 67 (62-71) | 27.5 (12.4-46.0) |
| mRNA-1273 | Alpha (B.1.1.7) | - | Qatar: 100.0 (91.8-100) | 90.9 (87.2-93.7) |
| mRNA-1273 | Beta (B.1.351) | - | Qatar: 96.4 (91.9-98.7) | 61.6 (55.3-67.5) |
| mRNA-1273 | Delta (B.1.617.2) |  | Qatar: 86.1 (78.0-91.3) | 71.4 (62.3-79.1) |
| Against COVID-19 symptomatic cases | | | | |
| Ad26.COV2.S | Beta (B.1.351) | South Africa: 52.0 (30.3-67.4) |  | 40.4 (25.5-56.3) |
| BNT162b2 | Alpha (B.1.1.7) | - | Canada: 89 (87-91)  Scotland: 92  United Kingdom: 93.7 (91.6-95.3) | 94.3 (87.9-97.5) |
| BNT162b2 | Delta (B.1.617.2) | - | Canada: 85 (59-94)  Scotland: 83  United Kingdom: 88.0 (85.3-90.1)  United Kingdom: 86 (83-88) | 95.1 (88.4-98.1) |
|  |  |  | Qatar: 56.1 (41.4-67.2) |  |
| mRNA-1273 | Alpha (B.1.1.7) | - | Canada: 91 (84-95) | 94.9 (86.8-98.2) |
| mRNA-1273 | Delta (B.1.617.2) | - | Canada: 72 (57-82)  Qatar: 85.8 (70.6-93.9) | 80.8 (60.7-92.3) |
| ChAdOx1 nCoV-19 | Alpha (B.1.1.7) | United Kingdom: 70.4 (43.6-84.5) | Canada: 75 (-98-97)  Scotland: 81  United Kingdom: 74.5 (68.4-79.4) | 76.3 (54.4-89.0) |
| ChAdOx1 nCoV-19 | Beta (B.1.351) | South Africa: 10.4 (-76.8-54.8) | - | 40.7 (19.5-62.3) |
| ChAdOx1 nCoV-19 | Delta (B.1.617.2) | - | Canada: 70 (46-83)  Scotland: 61  United Kingdom: 67.0 (61.3-71.8)  United Kingdom: 70 (66-74) | 41.1 (19.8-62.8) |
| ChAdOx1 nCoV-19 | Gamma (P.1) | - | Brazil: 77.9 (69.2-84.2) | 63.3 (38.3-81.2) |
| NVX-CoV2373 | Alpha (B.1.1.7) | United Kingdom: 86.3 (71.3-93.5) |  | 92.0 (43.7-99.4) |
| NVX-CoV2373 | Beta (B.1.351) | South Africa: 60.1 (19.9-80.1) |  | 54.4 (8.1-91.2) |
| Against severe COVID-19 | | | | |
| Ad26.COV2.S | Beta (B.1.351) | South Africa: 73.1 (40.0-89.4) | - | 52.5 (22.4-78.5) |
| BNT162b2 | Alpha (B.1.1.7) | - | Qatar: 100.0 (81.7-100.0) | 72.6 (0.1-99.5) |
| BNT162b2 | Beta (B.1.351) | - | Qatar: 100.0 (73.7-100.0) | 40.3 (0.0-97.2) |
| BNT162b2 | Delta (B.1.617.2) | - | Qatar: 97.3 (84.4-95.5) | 75.3 (0.1-99.6) |
| ChAdOx1 nCoV-19 | Gamma (P.1) | - | Brazil: 96.5 (81.7-99.3) | 99.8 (99.8-99.9) |
| NVX-CoV2373 | Alpha (B.1.1.7) | United Kingdom: 100 | - | 99.7 (99.5-99.9) |

### Supplementary figures

#### Figure S1. Predicted efficacy against infection for the variants of concern

Predicted efficacy against SARS-CoV-2 infection among reference lineages (blue bar), Alpha variant (orange bar), Beta variant (red bar), Gamma variant (green bar), and Delta variant (purple bar) were determined in **A)** mRNA-1273 and **B)** ChAdOx1 nCoV-19. The error bar represents the uncertainty interval.

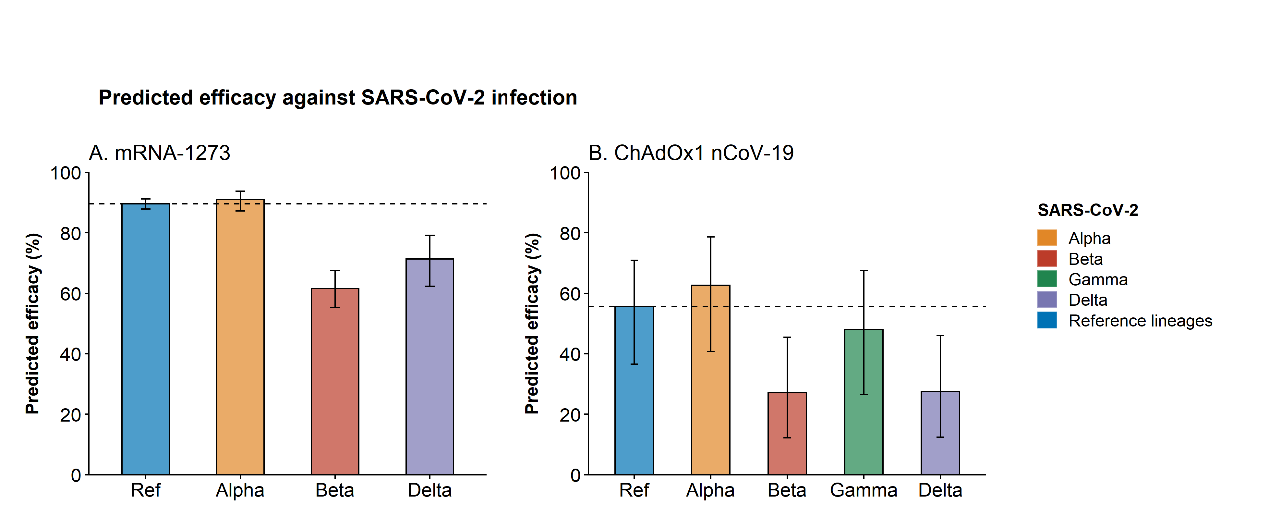

#### Figure S2. Predicted age-specific efficacy against symptomatic illness for the Delta variant for BNT162b2

The error bar represents the uncertainty interval.

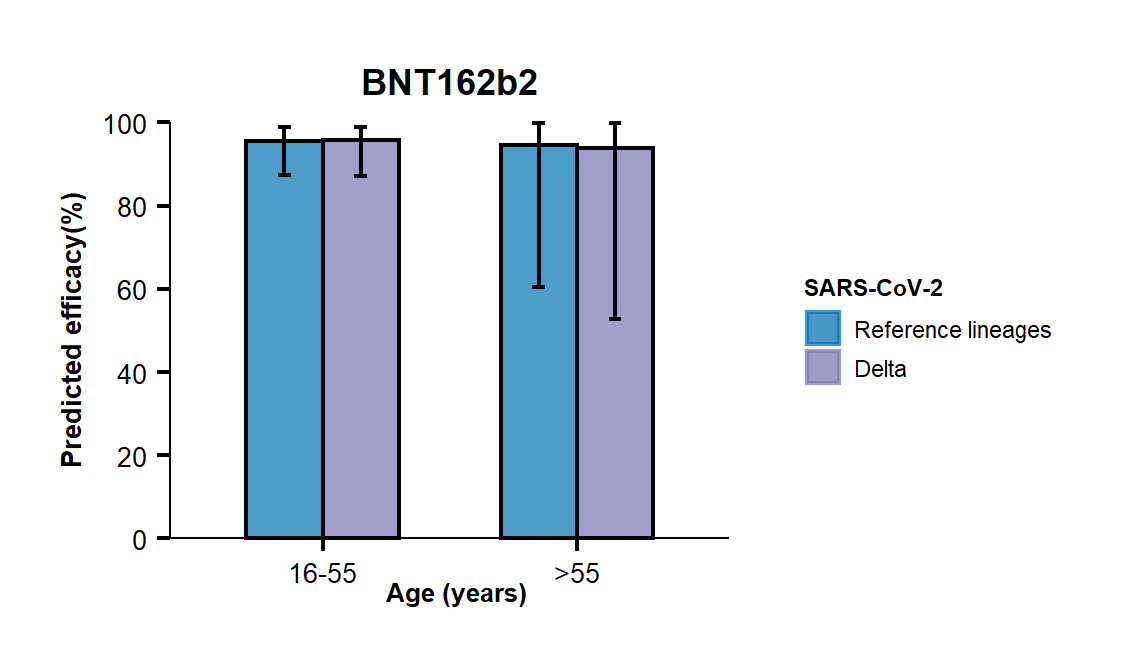

#### Figure S3. Predicted efficacy against symptomatic illness for the variants of concern

Predicted efficacy against symptomatic illness among reference lineages (blue bar), Alpha variant (orange bar), Beta variant (red bar), Gamma variant (green bar), and Delta variant (purple bar) were determined in **A)** BNT162b2, **B)** mRNA-1273, **C)** Ad26.COV2.S, **D)** ChAdOx1 nCoV-19, **E)** Gam-COVID-Vac and **F)** NVX-CoV2373. The error bar represents the uncertainty interval.

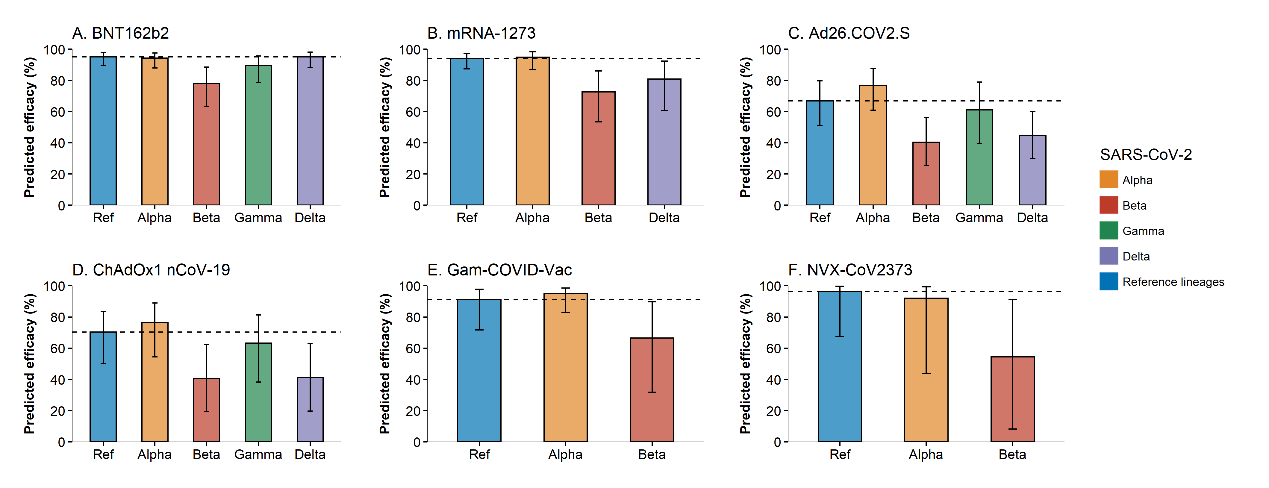

#### Figure S4. Predicted age-specific efficacy against the symptomatic illness for the variants of concern

Predicted age-specific efficacy against symptomatic illness among reference lineages (blue bar), Alpha variant (orange bar), Beta variant (red bar), Gamma variant (green bar) and Delta variant (purple bar) were determined in **A)** BNT162b2 and **B)** mRNA-1273. The error bar represents the uncertainty interval.

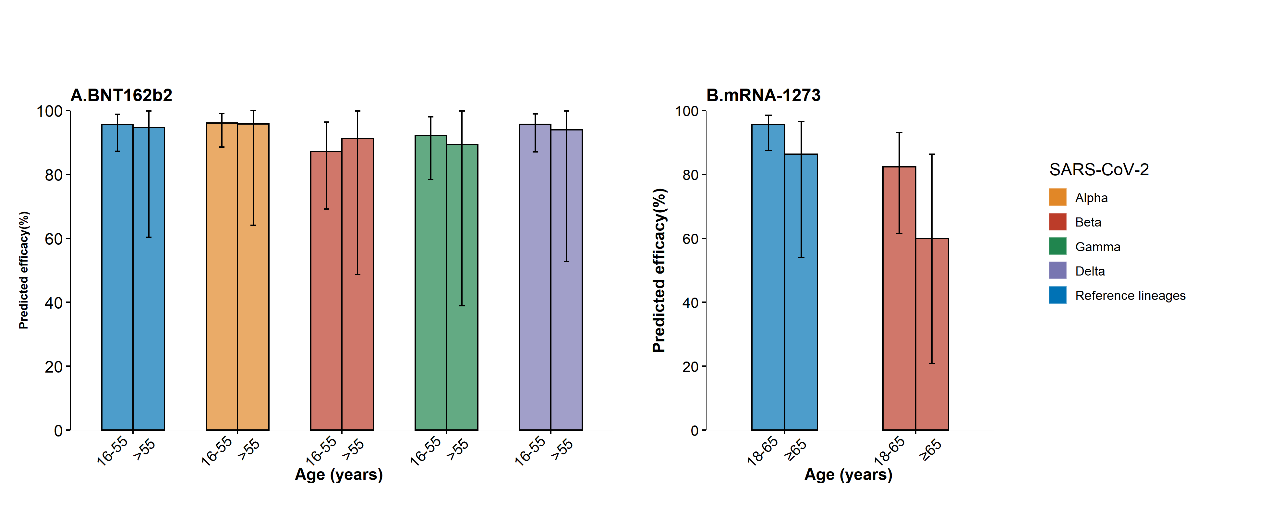

#### Figure S5. Predicted efficacy against the severe illness for the variants of concern

Predicted efficacy against severe illness among reference lineages (blue bar), Alpha variant (orange bar), Beta variant (red bar), Gamma variant (green bar) and Delta variant (purple bar) were determined in **A)** BNT162b2, **B)** mRNA-1273, **C)** Ad26.COV2.S and **D)** Gam-COVID-Vac. The error bar represents the uncertainty interval.

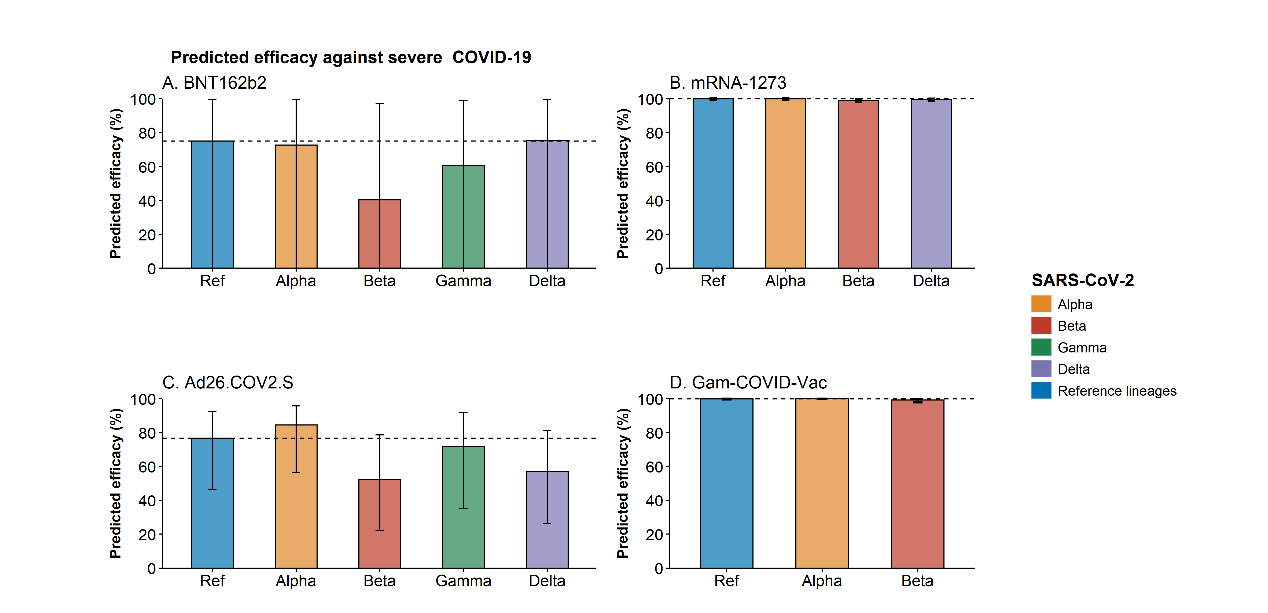

#### Figure S6. Comparison of predicted efficacy and observed efficacy or effectiveness across different endpoints for the variants of concern

Predicted efficacy was compared with observed efficacy and effectiveness against **A)** SARS-CoV-2 infection, **B)** symptomatic cases and **C)** severe COVID-19. The colors represent different SARS-CoV-2 variants, including Alpha (orange), Beta (red), Gamma (green) and Delta variant (purple bar). The error bar represents the uncertainty interval.

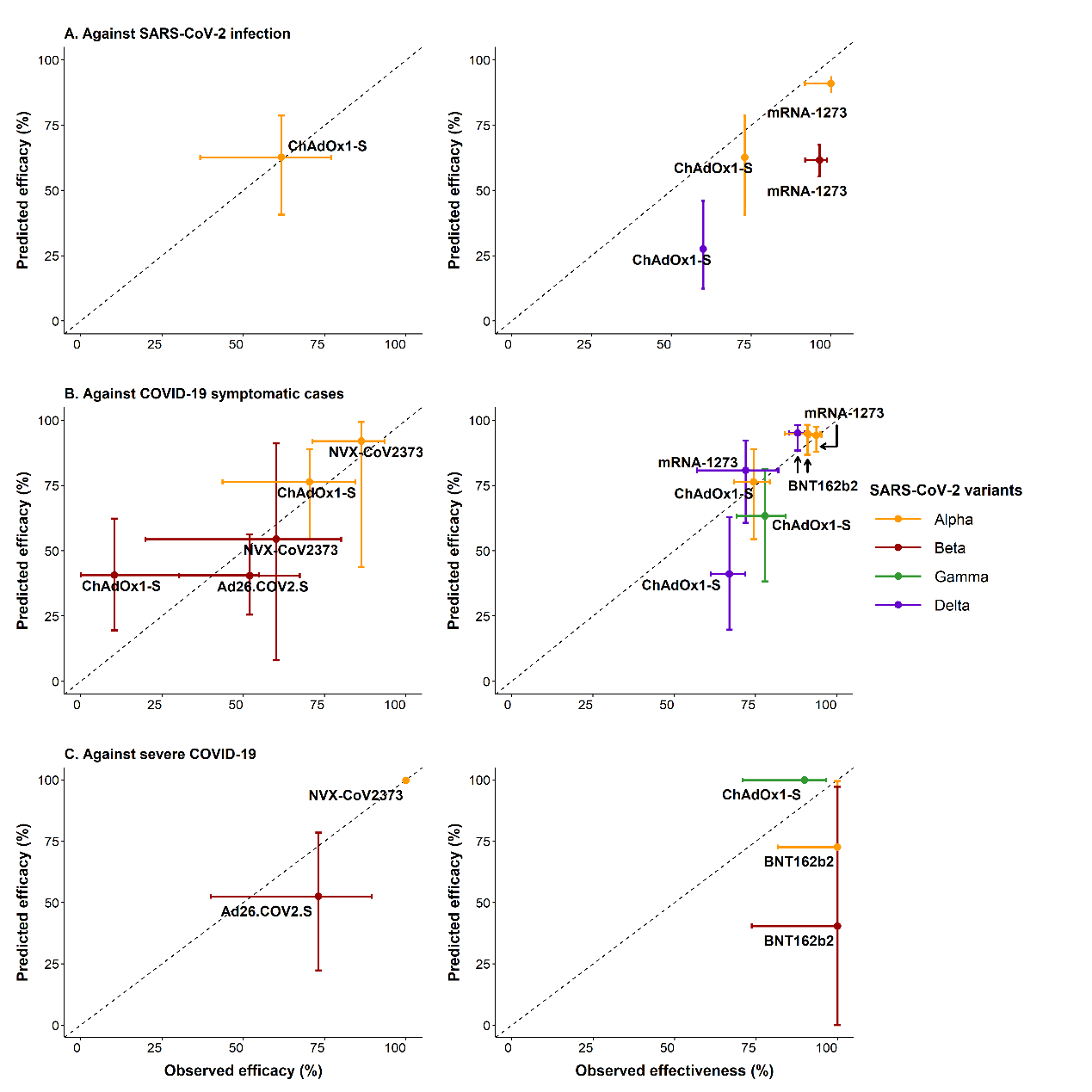
